# Predictive Modeling for Diabetes Using GraphLIME

**DOI:** 10.1101/2024.03.14.24304281

**Authors:** Flavia Costi, Darian Onchis, Eduard Hogea, Codruta Istin

## Abstract

The purpose of this paper is to present a detailed investigation of the advantages of employing GraphLIME (Local Interpretable Model Explanations for Graph Neural Networks) for the trustworthy prediction of diabetes mellitus. Our pursuit involves identifying the strengths of GraphLIME combined with the attention-mechanism over the standard coupling of deep learning neural networks with the original LIME method. The system build this way, provided us a proficient method for extracting the most relevant features and applying the attention mechanism exclusively to those features. We have closely monitored the performance metrics of the two approaches and conducted a comparative analysis. Leveraging attention mechanisms, we have achieved an accuracy of 92.6% for the addressed problem. The model’s performance is meticulously demonstrated throughout the study, and the results are furthermore evaluated using the Receiver Operating Characteristic (ROC) curve. By implementing this technique on a dataset of 768 patients diagnosed with or without diabetes mellitus, we have successfully boosted the model’s performance by over 18%.

## I. Introduction

In this paper, we explore the potential of advanced machine learning techniques in the early detection of diabetes. This chronic metabolic disorder, characterized by high blood sugar levels, has become a burgeoning health crisis globally. The escalating prevalence of diabetes underscores the necessity for innovative diagnostic methodologies. Our focus is on leveraging an attention mechanism within the framework of Graph Neural Networks (GNNs), enhanced by LIME, to identify individuals with or without diabetes [1].

Our approach employs an attention mechanism within machine learning, drawing inspiration from the selective focus aspect of human cognition. This mechanism enables the model to dynamically prioritize certain segments of the input data that are more relevant for accurate diagnosis. As can be seen in Table I, there are many more advantages that have led us to choose to use this technique. GNNs are adept at processing data in graph structures, a common form in medical data representation. LIME contributes by rendering these complex models more interpretable [20]. It approximates the GNN with a simpler, yet effective model, providing transparent and understandable explanations for each prediction. This aspect is important in medical diagnostics, as it not only provides insight into the model’s predictive behavior but also enhances trust and transparency in its outcomes, essential for healthcare practitioners. We want to explore the advantages of using our proposed method for predicting the presence or absence of diabetes in patients [2]. We have conducted a comprehensive comparison and analysis of the results obtained both before and after the implementation of our adapted method to evaluate the model’s performance. By employing our technique, we have successfully increased the accuracy of the predictions by over 18%.

**TABLE I.** Differences and Similarities between LIME and graphLIME.

| Feature | LIME | GraphLIME |
| --- | --- | --- |
| Specific for structured data |  | ✓ |
| Generalizable to any type of data | ✓ | ✓ |
| Provides local explanations | ✓ | ✓ |
| Suitable for graph-based models |  | ✓ |
| Simple and intuitive | ✓ |  |
| Manages complexity in graph data |  | ✓ |
| Versatile for various ML models | ✓ | ✓ |
| Detailed explanations for network structures |  | ✓ |
| Quick and straightforward explanations | ✓ | ✓ |

## II. Related work

The paper [4] is centered on developing machine learning models for diabetes prediction, with a strong emphasis on the use of explainable AI to improve the predictions’ trustworthiness. This research aims to offer a valuable tool for the early detection of diabetes, vital for effective management and treatment in healthcare. The method incorporates machine learning algorithms like decision trees, SVM, Random Forest, Logistic Regression, KNN, and ensemble techniques. Notably, the XGBoost classifier emerged as the most effective, complemented by the ADASYN technique to address class imbalance in medical datasets. The model demonstrated notable accuracy (81%), an F1 score of 0.81, and an AUC of 0.84, showcasing its reliability. Interpretability is achieved using LIME and SHAP, providing clarity on the model’s decision-making process. Our approach, inspired by authors of paper [4], diverges in utilizing GraphLIME instead of LIME. GraphLIME offers improved feature extraction and contextually rich insights, thereby enhancing model interpretations. By integrating an attention mechanism, we not only focus on key features but also add depth to the predictive analysis, making our approach distinct and more nuanced [4].

The paper [5] offers a detailed evaluation of various machine and deep learning techniques for Type 2 Diabetes prediction. It reviews a spectrum of models, assessing their effectiveness and pinpointing the most effective techniques for predictive modeling. The review notes the superior performance of tree-based algorithms and observes that, while deep neural networks are potent with large datasets, they often yield less optimal results. It underscores the significance of data balancing and feature selection in boosting model efficiency, with well-structured datasets leading to near-perfect outcomes. The article notably delves into the advantages of using graphs in deep learning, a key insight that influenced our decision to utilize GraphLIME over traditional LIME for feature extraction. Inspired by the article [11] we adapted its foundational concept to our dataset [7]. We further enhanced our methodology by integrating an attention mechanism, focusing on the relevant features identified using GraphLIME. This approach aimed to refine and improve upon the existing models for this widely used dataset, drawing on the systematic review’s findings and the pivotal role of GraphLIME [5]. The most relevant differences between LIME and GraphLIME can be observed in Table I.

## III. Proposed approach

### A. Purposes

Following the research studies presented in the section II, we noticed that explainability methods are not extensively explored. Most works stop at extracting the most relevant features, but they do not leverage this information to expand the developed model. However, we wanted to explore this field of explainability in this work and use the obtained information to enhance the model’s performance.

Building on the foundation laid by the studies presented in the section II, our secondary aim is to validate the hypothesis that employing graph-based approaches in explainability represents the most optimal strategy. Specifically, we aim to assess the effectiveness of using GraphLIME in extracting the most pertinent features from our dataset. This investigation is critical in demonstrating the practical utility and superiority of graph-based methods in elucidating complex data characteristics, thereby contributing to the advancement of explainability in machine learning [6].

### B. Objectives

The objective of our research is to implement and rigorously analyze the performance of the methodologies discussed earlier. We aim to make a substantial and positive contribution to the developed solution, enhancing and refining existing ideas to improve the overall performance of the model. Our aim is to observe how much using GraphLIME aids us in the development of the model, compared to the classical method of explainability, LIME. Through this analysis, we will be able to demonstrate the importance of using explainability methods in improving the model’s performance and, at the same time, highlight that leveraging this technique comes with benefits.

### C. Approach

For our experiments, we have used a public diabetes dataset from Kaggle [7] that comprises medical records of 768 patients (Fig. 1 to see how many patients suffer from diabetes and how many do not), categorized based on their diabetic condition. Eight of these columns contain independent data, encompassing various factors that contribute to the diagnosis of diabetes mellitus. The ninth column is dependent, indicating whether a patient has diabetes.

**Fig. 1.**
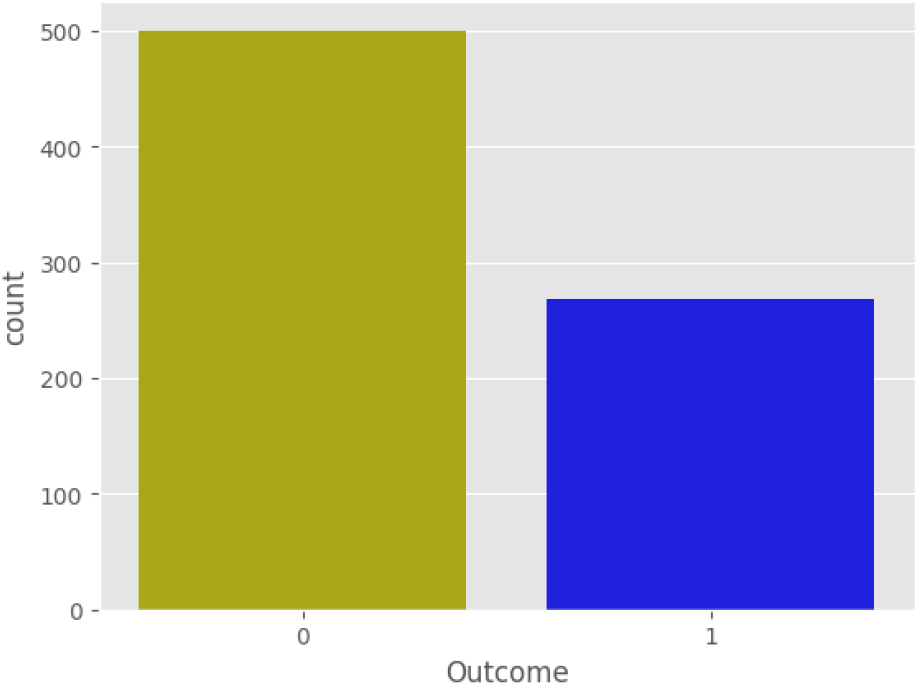
The number of patients with and without diabetes

As illustrated in the figure above (Fig. 1), the dataset encompasses 500 patients who do not have diabetes (indicated by a dependent feature value of 0) and over 250 patients who do have diabetes (with a dependent feature value of 1). This presents a disparity where there are approximately 50% fewer patients with the disease compared to those without it. Such an imbalance necessitates the balancing of the dataset. To balance our dataset, we used the SMOTE (Synthetic Minority Over-sampling Technique) technique [19]. This technique works by creating synthetic examples of the minority class, in our case, patients suffering from diabetes. This is achieved by interpolating between existing examples, contributing to the diversification and expansion of the dataset. By applying this, we were able to bring the number of cases in the minority class (patients with diabetes) to a level similar to the majority class (patients without diabetes).

To evaluate the performance of our model, we implemented the k-fold cross-validation method, dividing the dataset into 5 folds. This approach allowed us to obtain a more precise and robust estimation of our model’s effectiveness, avoiding potential biases that can arise when using a single dataset for testing. Through k-fold cross-validation, each example in the dataset had the chance to be used both in the training and testing sets. This helped us to verify if the model could generalize well on new data. In each of the 5 iterations of k-fold cross-validation, a different subset of data was chosen as the testing set, while the rest of the data were used for training the model [8].

The subsequent phase involves the development of a TensorFlow-based model, utilizing GNN. This step marks the transition from data preparation to the intricate process of model building, where we leverage the advanced capabilities of TensorFlow and the structural benefits of GNNs to extract meaningful insights and patterns from the dataset. This approach is expected to provide a comprehensive understanding of the data characteristics, aiding in the accurate identification and prediction of diabetic conditions among patients [9].

The schematic in Figure 2 provides a detailed overview for GNN process. This process begins with the utilization of a trained GNN and a series of graphs that are, ideally, similar in distribution to those used during the GNN’s training phase [10]. The method unfolds as follows:

**Fig. 2.**
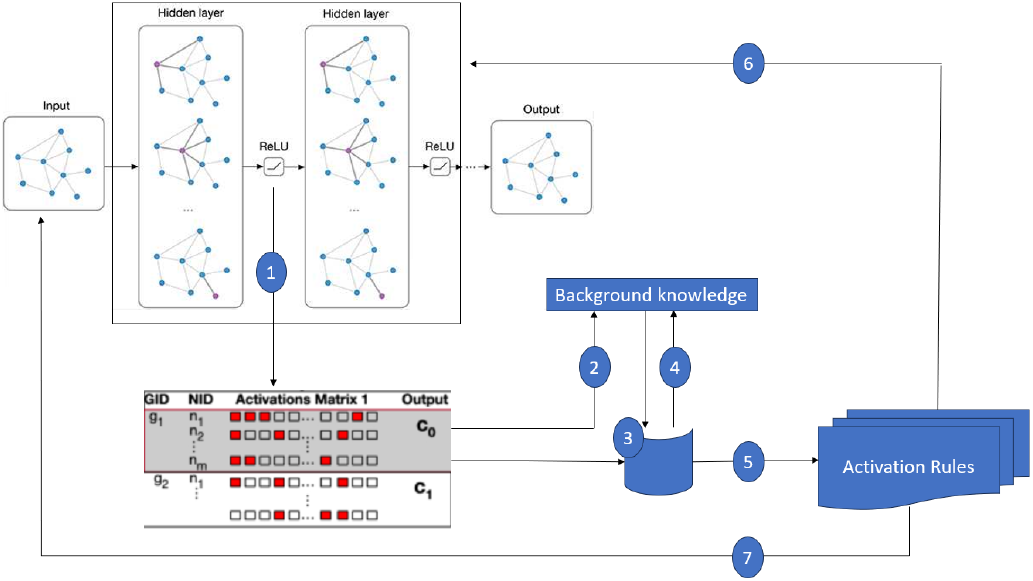
INSIDE-GNN Process: Sequential Discovery and Integration of Activation Rules for Enhanced Model Insight

1. *Construction of an Activation Matrix* We develop a binary matrix to encapsulate how the nodes within the graphs influence various vector components within the GNN. This matrix also correlates the nodes with the decisions derived from the GNN.
2. *Establishment of a Preliminary Model* This model serves as an initial framework, reflecting our base-level understanding of the data within the matrix. At this starting point, the model operates under the assumption that there is no inherent connection between the activations and the nodes in the graphs.
3. *The INSIDE-GNN Procedure* This step involves the application of the INSIDE-GNN process, which is designed to pinpoint the most significant rule of activation. It does this by analyzing both the activation matrix and the initial model.
4. *Model Refinement* Following the identification of a key activation rule, we update the initial model to integrate this new rule, thereby enriching our pattern set.
5. *Iterative Enhancement* We repeatedly cycle through steps 2 to 5, each time enhancing our model and pattern set. This process continues until it either yields no new significant rules or meets predefined criteria for cessation.
6. *Pattern-Based Explanations* Utilizing the collection of activation patterns, we generate explanations for each instance. This is achieved by implementing various strategies that involve node-based masks in alignment with the activation rules.
7. *Rule-Support Analysis* For each identified activation rule, we engage in an in-depth exploratory analysis, employing techniques like subgroup discovery in graph propositionalization or mining of subgraphs. The objective here is to conduct a detailed examination of the nodes that support these rules, thereby providing clear, interpretable insights into the aspects captured by the GNN.

Following the successful construction of our TensorFlow model, we embarked on a important phase: training the model. This step was pivotal in fine-tuning the model’s parameters to align with our dataset’s unique characteristics. Subsequent to the initial training, we initiated the generation of perturbed samples. Each of these samples underwent a meticulous training process, ensuring that our model was robust and capable of handling a variety of data alterations. The final stage in our methodology involved the computation of GraphLIME. This was a significant step, as GraphLIME plays an instrumental role in interpreting the model’s decisions by providing a clear and comprehensible explanation of the model’s behavior, particularly in relation to the perturbed samples. Through this comprehensive process, from model building to GraphLIME calculation, we aimed to create a model that is not only accurate but also transparent and interpretable in its decision-making. The Figure 3 shows all the mechanisms behind GraphLIME [11].

**Fig. 3.**
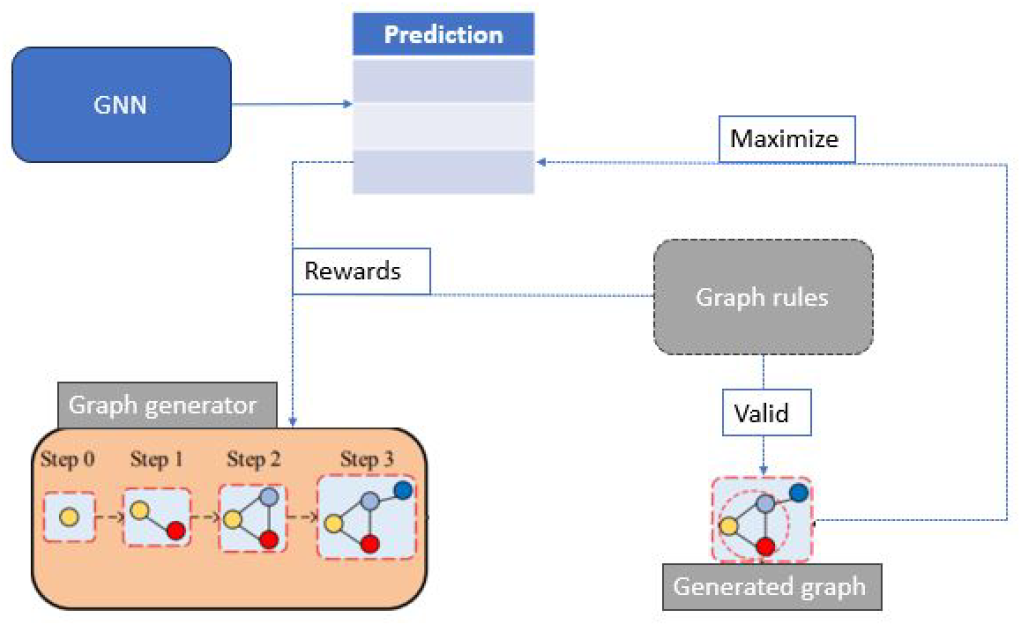
The architecture of GraphLIME

Upon successfully identifying the most pertinent features from our dataset, we proceeded to apply an attention mechanism, but exclusively to the extracted data. This focused application is designed to enhance our model’s precision and interpretability by concentrating on the key features. The outcomes of this mechanism are of significant interest, and we will thoroughly analyze and discuss the results in a subsequent section of our study. This analysis aims to shed light on the efficacy of the attention mechanism in refining model performance and providing deeper insights into the data.

## IV. Preliminary results

Our dataset comprises a moderate volume of data (details at III-C), a factor that favorably circumvented the challenges often associated with smaller datasets. Consequently, the solution we propose is ideally suited for datasets ranging from moderate to large in size. In cases involving smaller datasets, additional modifications would be necessary to adapt our approach.

The core objective of our solution is to meticulously identify and apply the most appropriate methods and techniques to yield accurate and reliable results. Our structure facilitates a detailed examination of factors leading to diabetes diagnosis, ensuring both thoroughness and precision in our analysis. We aim to use this data to create a model that is not only effective in diagnosing diabetes but also robust across different data volumes.

The resulting system that we envision is the one seen in Figure 4. The sequential workflow is demonstrated, starting with the diabetes dataset, followed by preprocessing steps and preparing the input graph for analysis. The target feature is highlighted and a toy example of feature relationships in the graph is shown. Subsequently, the preprocessed data is fed into our Graph Neural Network (GNN) architecture for processing. After obtaining the model’s output, the input graph and the we sample the neighbors of the target(red) node, we employ GraphLIME to derive post hoc feature importance, isolating those features with a positive impact on diabetes prediction. These significant features are then utilized within an attention mechanism to refine and focus the model’s predictive capabilities. Finally, the system generates its prediction, leveraging both the insights gained from feature importance analysis and the focused processing power of the attention mechanism.

**Fig. 4.**
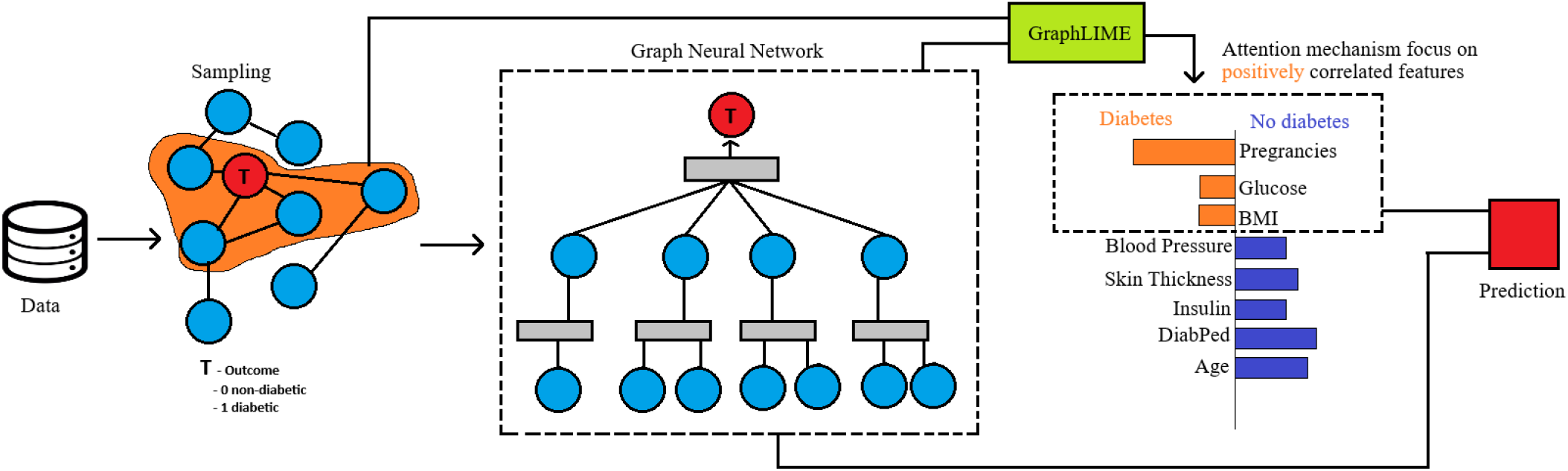
Full System Workflow Diagram. From dataset preprocessing to prediction, this figure outlines how our system utilizes GNN processing, GraphLIME for feature importance extraction, and key features with an attention mechanism for diabetes prediction.

### A. Designing and Evaluating a TensorFlow Model for Feature Importance in Diabetes Prediction

In order to effectively extract relevant features from our dataset, a two-step approach was initially employed. The first step involved splitting the dataset, a relevant process that set the stage for the subsequent model development. Once the dataset was appropriately segmented, we proceeded to construct a TensorFlow model, meticulously designed to optimize feature extraction.

The architecture of our TensorFlow attention model is structured around three Dense layers, each serving a specific role in the learning process. This specific choice is the result of extensive experimentation on a subset of the training data, where different combinations of hyperparameters and architecture shapes were tried. The final model contains in the first layer 64 units with the ‘relu’ activation function, the second layer includes 32 units, also with the ‘relu’ activation, and the final layer is composed of a single unit with the ‘sigmoid’ activation function. Using 64 units in the first layer offers a balance between the ability to process various shapes and complexities in the data and maintaining computational efficiency. A smaller number of units could have limited the model’s ability to learn complex features, while a larger number could lead to overfitting and reduced computational efficiency. The second layer with 32 units allows the model to further refine the features identified by the first layer, without adding unnecessary complexity. Choosing a final layer with a single unit and ‘sigmoid’ activation is relevant for binary classification tasks, providing a clear probabilistic output. Using a different function or more units in this layer could have impacted the model’s ability to provide precise binary predictions. Therefore, this specific configuration of Dense layers was determined by the need to balance the model’s learning capacity, avoiding overfitting and optimizing computational efficiency, while ensuring accuracy in binary classification tasks [14].

To train our model, we used the Adam optimizer, known for its effectiveness in deep learning and handling sparse gradients and noisy problems. This optimizer benefits from an adaptive learning rate and combines features from AdaGrad and RMSProp [15]. Our chosen loss function was binary cross-entropy, apt for binary classification as it measures the alignment of the model’s probability-based predictions with actual labels. Accuracy was our performance metric, indicating the proportion of correct predictions in binary classification.

An important step in our analysis involved calculating the GraphLIME explanation after generating perturbed samples and recording the model’s predictions. GraphLIME’s local interpretable explanations provide deep insights into the model’s data processing, important for ensuring its reliability and robustness, particularly with perturbed samples. Figure 5 highlights the significant features in our Kaggle dataset for predicting diabetes in women, with Pregnancies, Glucose, and BMI (Body Mass Index) being the most impactful.

**Fig. 5.**
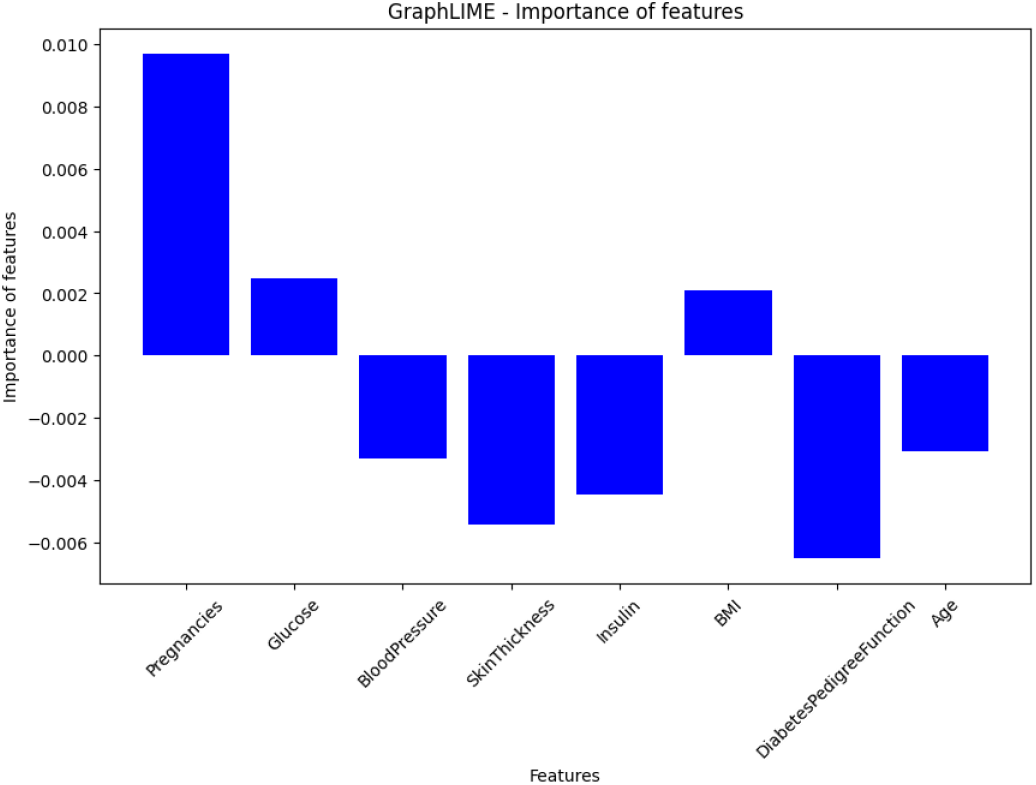
The importance of each characteristic in the prediction

### B. Application of the attention mechanism

In the realm of predictive modeling, attention mechanisms have emerged as a pivotal tool, primarily due to their capacity to enhance model interpretability and efficiency. The core objective of an attention mechanism is to enable the model to focus selectively on parts of the input that are most pertinent for a specific task, thereby improving the overall accuracy and interpretability of the model [16]. This selective focus is especially beneficial in disease prediction, where discerning subtle patterns and correlations within complex datasets is important [17].

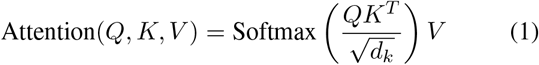

Where:

- *Q* represents the query matrix,
- *K* represents the key matrix,
- *V* represents the value matrix,
- *d*_*k*_ is the dimension of the keys and queries,
- The Softmax function is applied to normalize the attention scores.

The equation 1 expresses the core principles of the scaled dot-product attention mechanism, a pivotal component in advanced neural network architectures. This mechanism is particularly renowned for its ability to enhance both the interpretability and efficiency of models, especially in fields that require nuanced understanding of complex data, such as natural language processing or predictive modeling in healthcare. The equation operates on three fundamental components: the Query (*Q*), Key (*K*), and Value (*V*). These components play an important role in guiding the model’s focus to the most pertinent aspects of the input data. The process initiates with the calculation of the dot product between the Query and Key matrices. This step is essential, as it forms the basis for determining how each element of the Query aligns or correlates with the elements in the Key. Following this, the results from the dot product are meticulously scaled down by dividing them by the square root of the Key’s dimension 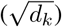. This relevant scaling step ensures that the softmax function, which follows, operates within an optimal range. It prevents the softmax from encountering extremely small gradients, which can be a significant hindrance in the training phase of a model, ensuring that the learning process remains stable and effective. The culmination of this process is the creation of a weighted sum of the Value components [18].

We incorporated an attention mechanism within our diabetes dataset to harness these advantages. Specifically, we employed two types of layers—ReLU and Sigmoid—to facilitate this process. ReLU was chosen for its ability to introduce non-linearity into the model without affecting the receptive fields of the convolution layer. On the other hand, the Sigmoid function was utilized due to its efficacy in binary classification tasks, which is a fundamental aspect of disease prediction. This combination of layers, in tandem with the attention mechanism, significantly contributed to the model’s performance. This integration resulted in a substantial increase in the model’s accuracy, achieving approximately 92.6%. To fully appreciate the outcome derived from the application of the two aforementioned techniques, it is essential to turn our attention to the following section. This segment meticulously delineates the results, offering a comprehensive understanding of how these methodologies converge to yield the observed findings.

## V. Obtained results

In this section, our focus shifts to exploring the potential of the implemented algorithms and assessing the performance of our model both before and after the application of the techniques discussed thus far. This comparative analysis aims to shed light on the efficacy of these methodologies, providing a clear understanding of their impact on the overall performance and accuracy of the model.

As can be seen from Table II, the accuracy of the model increased upon implementing the GraphLIME explainability method. Furthermore, by integrating an attention mechanism for relevant features within our dataset, we were able to enhance the model’s accuracy by an additional approximate 6 percent. The targeted application of an attention mechanism solely on relevant features within our dataset is of paramount importance. This selective focus ensures that the model’s computational resources are efficiently utilized, honing in on the most impactful aspects of the data. By doing so, it not only enhances the model’s accuracy but also improves its interpretability. This approach mitigates the risk of overfitting to irrelevant features and helps in maintaining the model’s robustness, making it more reliable and effective in real-world applications. We chose to utilize GraphLIME over LIME due to its distinct advantages in handling graph-based data. GraphLIME excels in providing graph-specific interpretability, adeptly capturing and elucidating complex node relationships and interactions that are relevant in graph structures. It is uniquely equipped to manage the complexities and structural dependencies inherent in graph data, offering localized explanations at the node level, which are essential for models trained on such data. Furthermore, GraphLIME stands out in terms of scalability and efficiency, effectively handling large graphs and complex network structures, a task where LIME is less optimized [11].

**TABLE 2.** Results of the model with and without GraphLIME and Attention Mechanism.

| Result description | Accuracy |
| --- | --- |
| Before applying GraphLIME and Attention Mechanism | 74.9% |
| After applying LIME | 82.9% |
| After applying GraphLIME | 86.2% |
| After applying GraphLIME and Attention Mechanism | 92.6% |

As illustrated in Figure 6, the obtained ROC curve showcases the high performance of our developed model. The curve, tending towards 1, signifies an exceptionally good predictive capability, underscoring the model’s effectiveness in the task at hand. This graphical representation is a clear indicator of the model’s robustness and accuracy in predictions. The complete code for predicting diabetes in women using the model developed and detailed in this paper is available for reference and use at the following [link Github](https://github.com/cflavia/GraphLIME/tree/main).

**Fig. 6.**
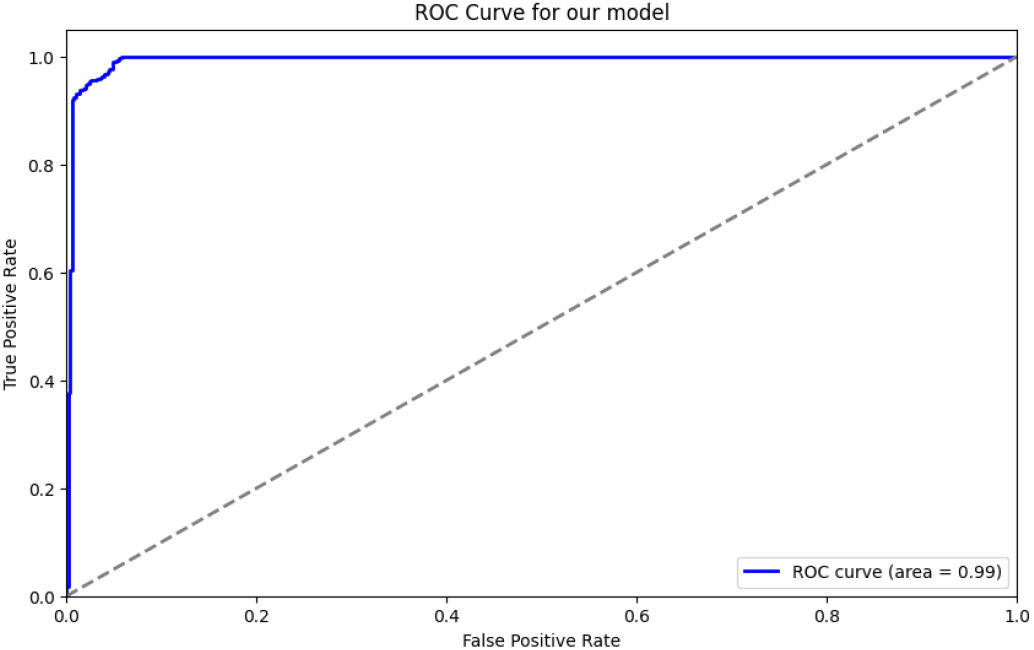
The ROC Curve for our developed model

## VI. Discussion

Through the application of our developed model, we have achieved a more precise and realistic prediction of diabetes based on the provided characteristics. The dataset utilized was of a moderate size in terms of patient numbers, and in this context, our model performed exceptionally well, surpassing expectations. The use of GraphLIME in diabetes prediction, as compared to LIME, has shown distinct advantages. Despite the limited information available on GraphLIME, we successfully developed a robust model capable of extracting relevant features. Leveraging these features, we incorporated an attention mechanism to enhance the predictability level, demonstrating the efficacy of our approach in diabetes prediction.

We successfully achieved a significant milestone in enhancing the accuracy of our predictions by approximately 18%. It was observed that the sole use of LIME or GraphLIME did not substantially increase the model’s accuracy. This led to the realization that the inclusion of a simple attention mechanism was necessary to achieve this level of improvement. The integration of this mechanism played an important role in fine-tuning our model’s performance, demonstrating that a combination of advanced techniques is often required to realize substantial gains in predictive accuracy.

As a future direction, we aim to adapt our solution to the multiclass case in order to observe the model’s performance in this context. Additionally, we intend to study the results obtained from implementing the model for the multi-class scenario and explore how we can enhance this technique to achieve predictions that closely align with reality [21]. Therefore, our objectives include the extension and in-depth exploration of this technique for multi-class applications.

## Data Availability

All data produced are available online at https://www.kaggle.com/datasets/mathchi/diabetes-data-set

## References

[1] T. F. Melo, E. J. F. Lima, R. A. Fagundes, R. M. Assunção, I. A. A. Araújo, D. J. M. Fagundes, M. R. S. Gadelha, “Machine learning and deep learning predictive models for type 2 diabetes: a systematic review,” Diabetol. Metab. Syndr., vol. 13, no. 1, 2021. https://dmsjournal.biomedcentral.com/articles/10.1186/s13098-021-00767-9

[2] Peng Mei, Yu hong Zhao, “Dynamic network link prediction with node representation learning from graph convolutional networks” Sci Rep 14, 538, 2024. 10.1038/s41598-023-50977-6

[3] Q. Lai, S. Khan, Y. Nie, H. Sun, J. Shen, and L. Shao, “Understanding more about human and machine attention in deep neural networks,” IEEE Transactions on Multimedia, vol. 23, pp. 2086–2099, 2020, IEEE.

[4] I. Tasin, T. U. Nabil, S. Islam, and R. Khan, “Diabetes prediction using machine learning and explainable AI techniques,” Healthcare Technology Letters, vol. 10, no. 1-2, pp. 1–10, 2023, Wiley Online Library.

[5] L. Fregoso-Aparicio, J. Noguez, L. Montesinos, and J. A. García-García, “Machine learning and deep learning predictive models for type 2 diabetes: a systematic review,” Diabetology & Metabolic Syndrome, vol. 13, no. 1, pp. 1–22, 2021, BioMed Central.

[6] C. Agarwal, O. Queen, H. Lakkaraju, and M. Zitnik, “Evaluating explainability for graph neural networks,” Scientific Data, vol. 10, no. 1, p. 144, 2023, Nature Publishing Group UK London.

[7] Kaggle. Diabetes Data Set. https://www.kaggle.com/datasets/mathchi/diabetes-data-set. [Accessed Date of Access]

[8] Trevor Hastie, Robert Tibshirani, and Jerome Friedman. The Elements of Statistical Learning: Data Mining, Inference, and Prediction. Springer Series in Statistics, 2009.

[9] TensorFlow. TensorFlow Graph Neural Networks. https://github.com/tensorflow/gnn, 2024. Accessed in January 2024.

[10] Luca Veyrin-Forrer, Ataollah Kamal, Stefan Duffner, and Celine Robardet. On GNN Explanability with Activation Rules. [Source publication name], October 2022. Available: https://www.researchgate.net/publication/364280740OnGNNexplanabilitywithactivationrules

[11] Qiang Huang, Makoto Yamada, Yuan Tian, Dinesh Singh, s i Yi Chang. Graphlime: Local interpretable model explanations for graph neural networks. IEEE Transactions on Knowledge and Data Engineering, 2022. IEEE.

[12] Abdulhakim Salum Hassan, I Malaserene, and A Anny Leema. Diabetes mellitus prediction using classification techniques. Int. J. Innov. Technol. Explor. Eng, 9(5):2080–2084, 2020.

[13] QuestionPro. (2024). Correlation Matrix: What is it, How It Works with Examples. Retrieved from https://www.questionpro.com/blog/correlation-matrix/

[14] S. Katakam, “Transfer Learning in TensorFlow: Feature Extraction,” Sandesh’s Machine Learning Journal, February 18, 2022. [Online]. Available: https://sandeshkatakam.github.io/My-Machinelearning-Blog/deep%20learning/neuralnetworks/tensorflow/transfer-learning/feature-extraction/2022/02/18/Transfer-Learning-in-TensorFlow-Feature-Extraction.html

[15] H. Kim, S. Lee, Y. Kim, and S. Hwang, “Classification and Prediction on the Effects of Nutritional Intake on Overweight/Obesity, Dyslipidemia, Hypertension and Type 2 Diabetes Mellitus Using Deep Learning Model: 4-7th Korea National Health and Nutrition Examination Survey,” Nutrients, vol. 13, no. 6, pp. 1958, 2021. [Online]. Available: https://pubmed.ncbi.nlm.nih.gov/34073854/

[16] Vaswani, A., Shazeer, N., Parmar, N., Uszkoreit, J., Jones, L., Gomez, A. N., … & Polosukhin, I. (2017). Attention is all you need. In Advances in neural information processing systems (pp. 30–48).

[17] G. Bansal, B. R. Singh, R. Kumar, and G. Kaur, “Enhancing heart disease prediction using a self-attention-based transformer model,” Scientific Reports, vol. 14, no. 1, pp. 1–12, 2024. [Online]. Available: https://www.nature.com/articles/s41598-024-51184-7

[18] S. Rome, “Understanding Attention in Neural Networks Mathematically,” 2018. [Online]. Available: http://srome.github.io/Understanding-Attention-in-Neural-Networks-Mathematically/

[19] Darian M. Onchis, Pavel Rajmic, Generalized Goertzel algorithm for computing the natural frequencies of cantilever beams, Signal Processing, Volume 96, Part A, 2014, Pages 45-50, ISSN 0165-1684, 10.1016/j.sigpro.2013.07.026.

[20] Secasan, C.C.; Onchis, D.; Bardan, R.; Cumpanas, A.; Novacescu, D.; Botoca, C.; Dema, A.; Sporea, I. Artificial Intelligence System for Predicting Prostate Cancer Lesions from Shear Wave Elastography Measurements. Curr. Oncol. 2022, 29, 4212–4223. 10.3390/curroncol29060336

[21] C. Istin, A. Doboli, D. Pescaru and H. Ciocarlie. Impact of coverage preservation techniques on prolonging the network lifetime in traffic surveillance applications, 2008 4th International Conference on Intelligent Computer Communication and Processing, Cluj-Napoca, Romania, 2008, pp. 201–206, doi: 10.1109/ICCP.2008.4648373.

